# Effect of Sinotubular Junction Size on TAVR Leaflet Thrombosis: A Fluid-structure Interaction Analysis

**DOI:** 10.1101/2023.11.13.23298476

**Authors:** David Oks, Symon Reza, Mariano Vázquez, Guillaume Houzeaux, Brandon Kovarovic, Cristóbal Samaniego, Danny Bluestein

**Author notes:** Corresponding author: Prof. Danny Bluestein, Department of Biomedical Engineering Stony Brook University, Stony Brook, NY 11794-8084, USA. These authors contributed equally and share the first authorship (David Oks, Symon Reza). **Competing Interests:** Author D.B. has an equity interest in Polynova Cardiovascular Inc. Author B.K. is a consultant of Polynova Cardiovascular Inc. Author M.V. is the CTO/CSO of ELEM Biotech S.L. Author D.O. is a Business Development Engineer at ELEM Biotech S.L. Author G.H. and C.S. have equity interests in ELEM Biotech S.L. The other authors declare that they have no competing interests.

## Abstract

**Purpose:** TAVR has emerged as a standard approach for treating severe aortic stenosis patients. However, it is associated with several clinical complications, including subclinical leaflet thrombosis characterized by Hypoattenuated Leaflet Thickening (HALT). A rigorous analysis of TAVR device thrombogenicity considering anatomical variations is essential for estimating this risk. Clinicians use the Sinotubular Junction (STJ) diameter for TAVR sizing, but there is a paucity of research on its influence on TAVR devices thrombogenicity.

**Methods:** A Medtronic Evolut® TAVR device was deployed in three patient models with varying STJ diameters (26, 30, and 34mm) to evaluate its impact on post-deployment hemodynamics and thrombogenicity, employing a novel computational framework combining prosthesis deployment and fluid-structure interaction analysis.

**Results:** The 30 mm STJ patient case exhibited the best hemodynamic performance: 5.94 *mmHg* mean transvalvular pressure gradient (TPG), 2.64 *cm*^2^ mean geometric orifice area (GOA), and the lowest mean residence time (T_R_) - indicating a reduced thrombogenic risk; 26 mm STJ exhibited a 10 % reduction in GOA and a 35% increase in mean TPG compared to the 30 mm STJ; 34 mm STJ depicted hemodynamics comparable to the 30 mm STJ, but with a 6% increase in T_R_ and elevated platelet stress accumulation.

**Conclusion:** A smaller STJ size impairs adequate expansion of the TAVR stent, which may lead to suboptimal hemodynamic performance. Conversely, a larger STJ size marginally enhances the hemodynamic performance but increases the risk of TAVR leaflet thrombosis. Such analysis can aid pre-procedural planning and minimize the risk of TAVR leaflet thrombosis.

## 1 Introduction

Calcific aortic valve disease (CAVD) is the most common valvular heart disease worldwide, resulting in over 182,000 aortic valve replacements each year in the U.S. alone [1]. Surgical Aortic Valve Replacement (SAVR) and Transcatheter Aortic Valve Replacement (TAVR) are two of the most commonly used procedures to treat CAVD patients. TAVR, a minimally invasive procedure was initially introduced to treat inoperable patients with severe aortic stenosis [2]. It has become the standard of care for high- and intermediate-risk patients demonstrating comparable performance to SAVR [3–5]. Recently, TAVR has also been extended to younger patients [6, 7]. Despite its fast adoption, the widespread use of TAVR is hampered by the evidence of post-TAVR subclinical leaflet thrombosis [8–10], which has been suggested as the underlying reason for HALT, leading to limited leaflet motion [11, 12]. Long-term outcome studies have revealed that TAVR leaflet degeneration and thrombosis are increasing in number and pose a significant risk to young and low-risk TAVR recipients [4, 5]. Recently, clinical studies have found evidence of early leaflet thrombosis formation [5] and that is associated with an earlier risk of structural valve degeneration. Hence, durability remains one of the major concerns for TAVR technology. Therefore, it is important to thoroughly investigate the thrombogenic risk of TAVR devices.

While planning interventions, clinicians consider patient-specific parameters to choose the optimal device for the patient. Previous studies analyzed the performance of TAVRs with respect to some established anatomical parameters, such as, the aortic annulus diameter and eccentricity, degree of stenosis, level of calcification in leaflets, or length and diameter of the Sinus of Valsalva (SoV). The size of the Sinotubular Junction (STJ) (see Figure 1[13]) has not been thoroughly investigated to determine its influence on post-TAVR outcomes. Supra-annular TAVR devices primarily anchor at the annular region with the calcified native leaflets. The crown region of the stent anchors in proximity to the STJ region that acts as the secondary anchorage zone. The diameter of the STJ affects the eccentricity of the stent at the crown region. Since the leaflet ends are stitched to the stent close to the crown region, it is likely that the leaflet dynamics will be affected by the changes in STJ diameter. This may lead to inferior hemodynamic performance.

**Figure 1:**
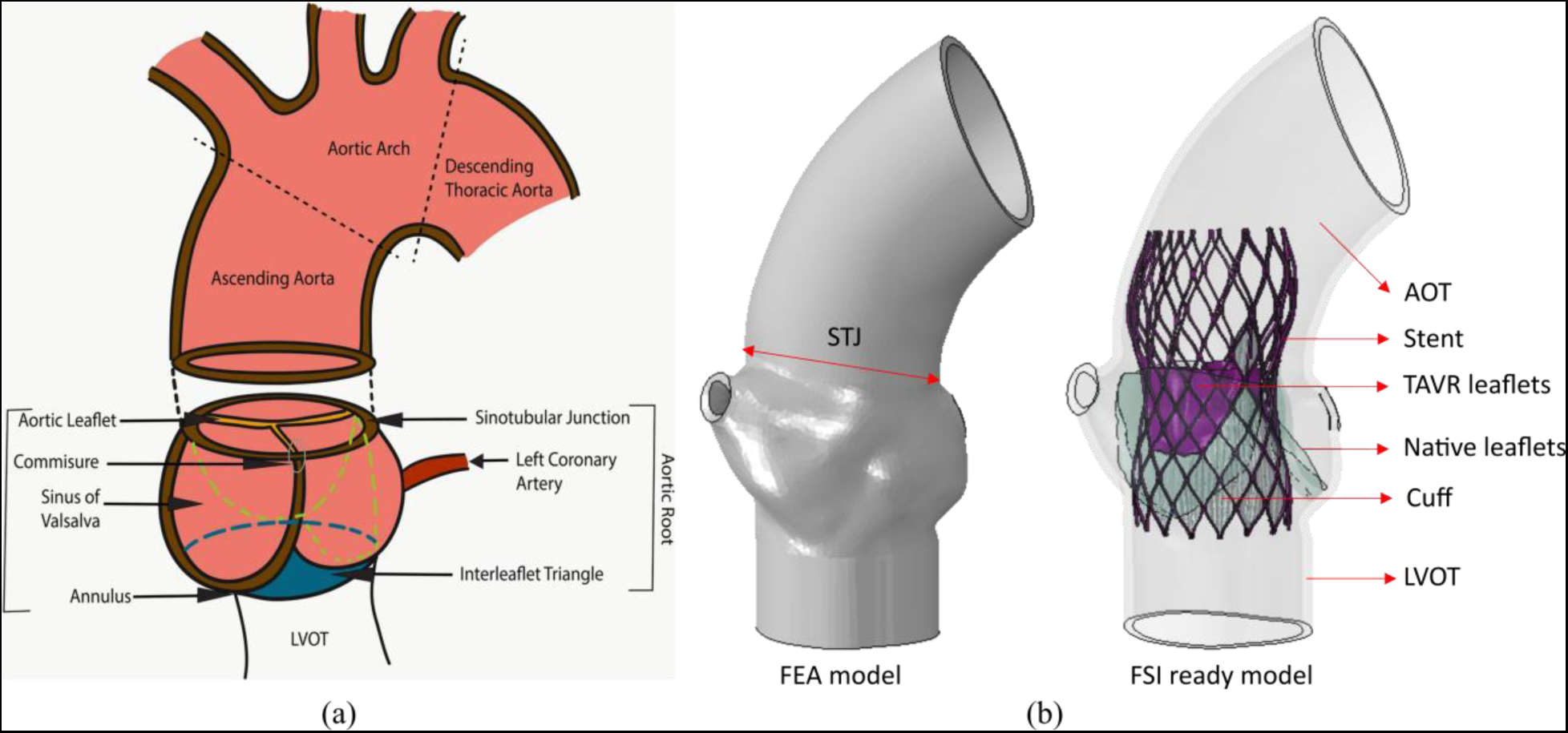
(a) Anatomy of the aortic root [13] (reproduced under Creative Commons Attribution 4.0 International License). (b) The FEA model (left) and the FSI ready model with the deployed TAVR device (right).

Previously, a clinical study by Nappi et al. has indicated that the dynamic variation of the STJ diameter during the cardiac cycle may play an important role in the risk of leaflet thrombosis and degeneration post-TAVR [14]. The STJ diameter has also been related to coronary obstruction, reporting a low STJ-to-TAVR diameter ratio to be a predictor of left main trunk obstruction [15]. The available *in-silico* studies that analyze the effect of STJ on native aortic valve performance are focused on regurgitation, tissue mechanics, and standard metrics of AVR performance. Pan et al. used FSI simulations to study the effect of STJ and sinus diameters on native aortic valve closure to prevent regurgitation [16]. Marom et al. performed FSI simulations to determine the influences of aortic annulus diameter and STJ-to-aortic annulus diameter ratio on native aortic valve hemodynamics and tissue mechanics and to suggest optimal values [17]. However, the effect of the STJ parameters on the TAVR deployment and the resulting hemodynamic performance of TAVR has not been studied. Therefore, analyzing the effect of STJ size on the deployment, hemodynamic performance, and thrombogenic risk of TAVR devices, may provide crucial information to optimize device selection. Consequently, this analysis may help reduce the risk of leaflet thrombosis and degeneration in the long run.

Thrombosis in prosthetic heart valves may be initiated by contact activation as well as by exposing platelets, the principal cell initiating blood clotting, to elevated shear stresses and recirculation flow patterns. When the cumulative effect of shear stress and exposure time is above a certain threshold, platelets are more likely to activate, triggering the coagulation cascade [18–20]. Low flow rate, regions of flow stasis and stagnation near the valve have been suggested as the source of leaflet thrombosis since those have been observed in *in-vitro* and *in-silico* studies [21–26]. Identifying regions of long residence times, high shear, and elevated platelet stresses is crucial to assess the thrombogenic potential of bioprosthetic aortic valves and improve their designs. Nonetheless, determining these parameters in experimental studies or clinical measurements has proven extremely challenging. We developed a Device Thrombogenicity Emulation (DTE) methodology [27, 28] based on flow-induced stress accumulation on platelets that accounts for the combined effect of stress and exposure time. It has been validated and used to analyze the thrombogenic risk associated with various cardiovascular devices, such as, Mechanical Heart Valves (MHVs) [29–31], Mechanical Circulatory Support (MCS) devices [28, 32] and TAVR devices [22, 33–35]. A predictive model for clinical and subclinical leaflet thrombosis has been proposed recently, based on Eulerian flow characteristics such as the percentage of stasis volume and average Wall Shear Stress [36, 37]. A computational study that additionally incorporates the Lagrangian flow-induced Stress Accumulation (SA) into this Eulerian framework enables a comprehensive assessment of device thrombogenic risk.

The current work studies the effect of the patient’s STJ diameter on standard metrics of valve prosthesis performance [38], and the associated thrombogenic risk. Our study addresses the following questions:

1. How does the STJ diameter affect the structural implantation and hemodynamic performance of TAVR as evaluated through the measurements of Transvalvular Pressure Gradient (TPG) and Geometric Orifice Area (GOA) in comparison to standard values of reference?
2. What is the effect of the STJ diameter on the thrombogenic risk in TAVR patients as indicated by Stress Accumulation (SA) on the platelets and flow characteristics, e.g., Residence Time (*T_R_*), Wall Shear Stress (WSS)?

## 2 Methods

The current work consists of two steps: the first uses structural FEA to simulate the deployment of TAVR in reconstructed patient-specific aortic roots, including the modeling of the Neosinus cavity formed post deployment. The second step uses the FEA simulations of the patient-specific geometries to solve the FSI between blood flow and the TAVR device’s leaflets.

### 2.1 Model reconstruction

A patient with 23.5mm average CT-derived annular diameter and 30 mm STJ diameter was selected for this study. The anonymized patient’s CT scan was acquired at Stony Brook University Hospital under the approval of the local institutional review board. The patient model, shown in Figure 2a, was reconstructed following the procedures described in our previous study [39]. Briefly, the pre-TAVR CT scan of the selected patient was imported as DICOM file and segmented into 3D surface meshes using *ITK-SNAP 3.6* [40]. The leaflets were reconstructed through surface interpolation, following the manually extracted coordinates of aortic leaflet landmarks. The generated surfaces were then meshed using *ANSYS SpaceClaim* and *Fluent Meshing tools* (ANSYS Inc., Canonsburg, PA) [34]. The meshed surface aortic sinus was then created in *Abaqus CAE* (SIMULIA, Dassault Syst**è**ms, Providence, RI) [41] to incorporate the wall thickness. Previously, clinical studies have analyzed the STJ cover index (STJCI), measured by the ratio between the CT-derived average STJ diameter and the nominal prosthesis crown diameter, as a parameter for analyzing the TAVR outcome [42]. To study the effect of STJ size, the concept of clinically driven virtual patient cohort that focuses on targeting sub-populations for specific applications was adopted [43]. In this study, the patient specific STJ size was parametrically adjusted while keeping the aortic annulus and the left ventricle outflow tract (LVOT) unchanged (see Figure 2a), resulting in three models with STJ diameters of *d*^*p*^= 26, 30, and 34 mm. The sizes were chosen based on literature where the STJ size of male TAVR patients is in the range of 29.8 ± 4.2mm [44]. The STJCI are therefore 0.82, 0.94, and 1.06 for the patients with STJ diameters of 26, 30, and 34 mm, respectively.

**Figure 2:**
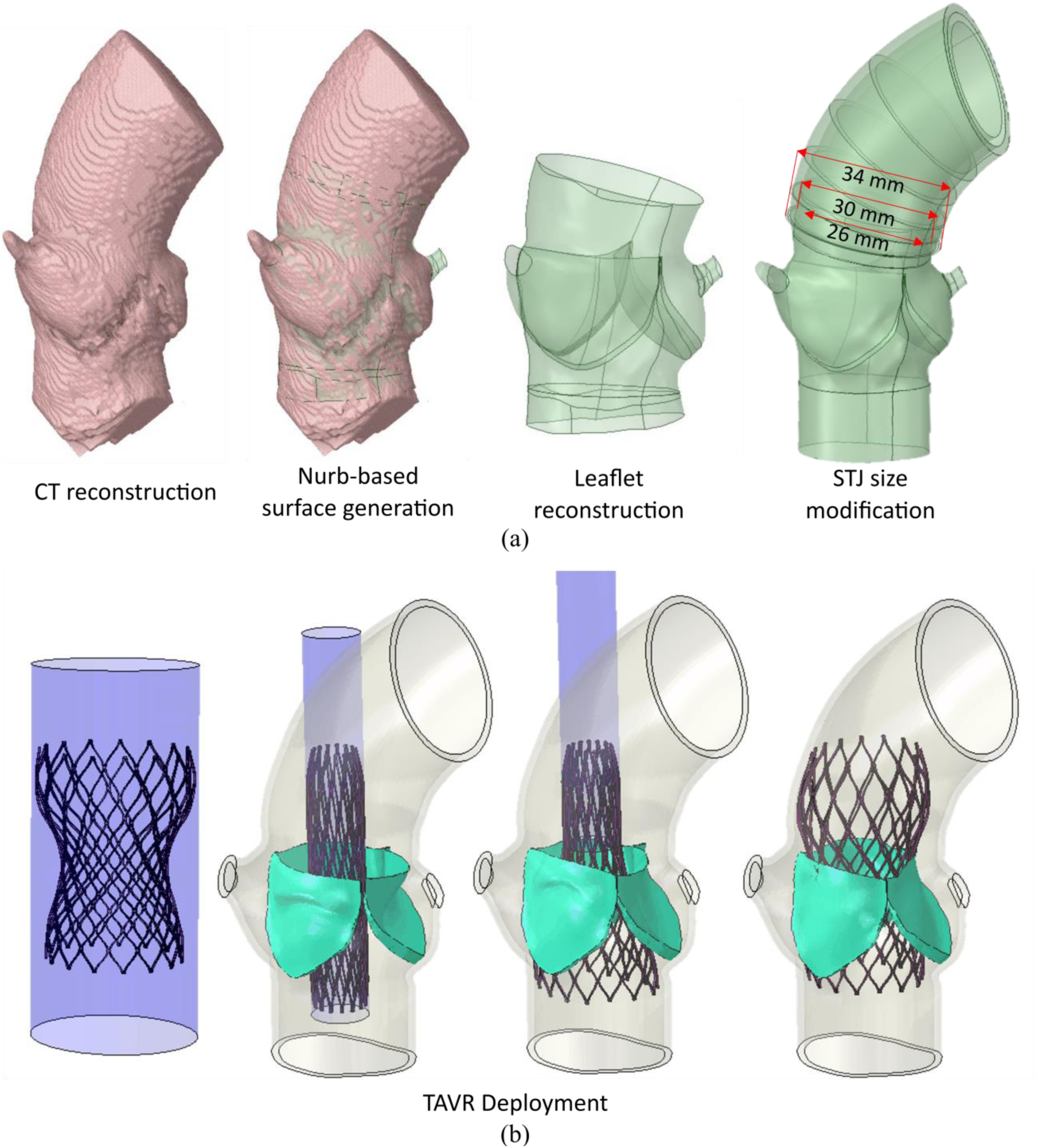
(a) Illustration of model reconstruction steps from CT and the modification of STJ diameter, (b) Illustration of uncrimped stent and crimper, crimped stent placed inside the patient aortic annulus model, TAVR device deployment, and TAVR device deployed inside the patient model with an STJ diameter of 26mm (left to right). The native valve which is pressed against the sinus wall during deployment is shown in green.

### 2.2 TAVR deployment modeling

With the abovementioned anatomical features, *Medtronic* Evolut^®^ 26 mm TAVR device was chosen and deployed in each one of the three anatomical models Figure 2b. The implantation depth was chosen to be 4mm following the recommended implantation depth for the Medtronic Evolut^®^ 26mm TAVR device (3−5 mm). The crimping and deployment steps were simulated using *Abaqus Explicit 2019* [41]. The stent was modeled as a superelastic Nitinol alloy (14-constants VUMAT material available in *Abaqus*) [45]. Dedicated Ogden 3^rd^-degree isotropic hyperelastic material models calibrated with biaxial test measurements were used for the aortic wall and each leaflet correspondingly [46, 47]. The prosthetic leaflets were then mapped to the deformed TAVR device for further hemodynamic study. Commissural alignment was ensured during the deployment process. Detailed information on the TAVR crimping and deployment simulations can be found in our previous studies [22, 33, 34, 48]. Subsequently, the resultant geometries of the deployed systems were exported as STLs, which were in turn used to construct the boundary and volume meshes with *ANSA BETA CAE* [49], with dimensions specified in Table 1. Linear tetrahedra were used to discretize the fluid volumes, while a hybrid linear hexahedron pentahedron mesh was used for the prosthesis leaflets. These meshes were then used for the FSI simulations described in the next section. A mesh convergence analysis for an analogous setup, demonstrating mesh independence for the aortic valve FSI problem can be found in [50].

**Table 1:**
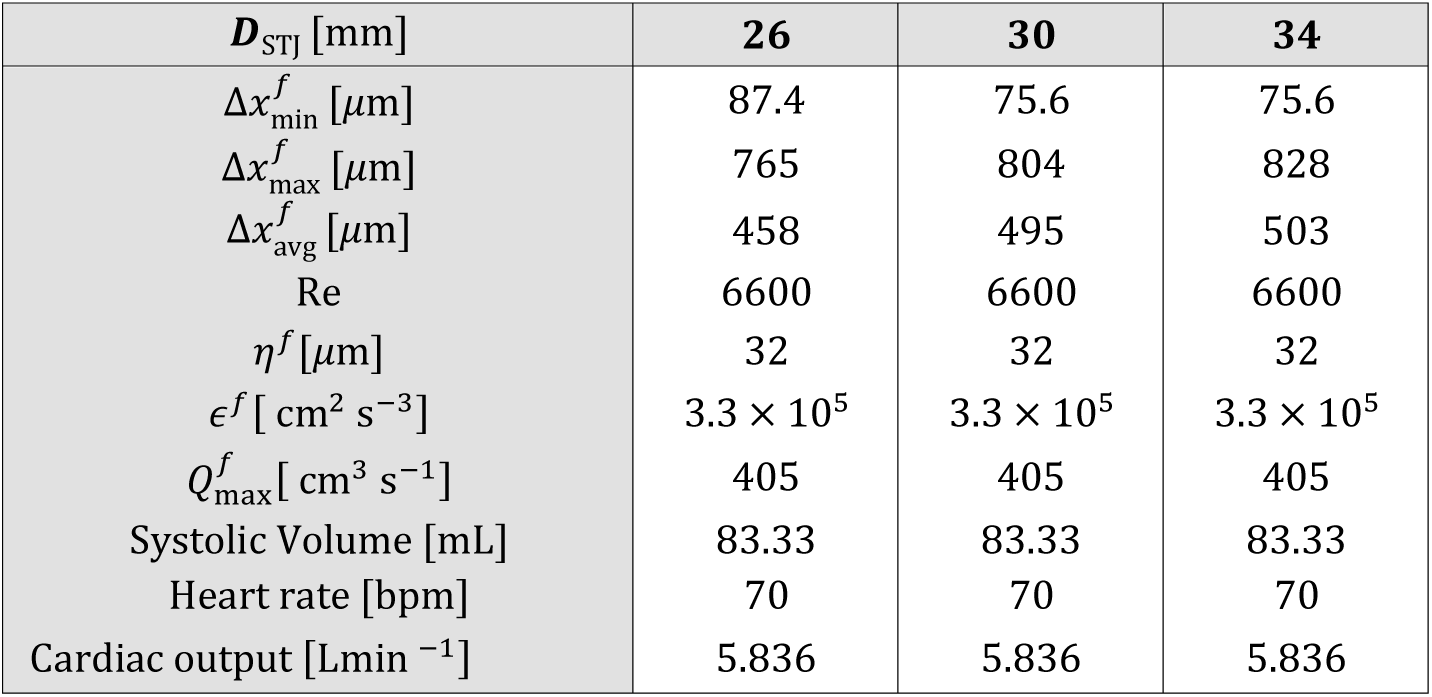
Mesh and flow characteristics, for each average STJ diameter, ***D***_*STJ*_, where *Δx*^*f*^_*min*_, *Δx*^*f*^_*max*_, and *Δx*^*f*^_*avg*_ are the minimum, maximum, and average element sizes respectively. The Reynolds number is *Re, η^f^* is the Kolmogorov microscale, *ϵ*^*f*^ is the average rate of viscous dissipation, *Q*^*f*^_*max*_ the maximum flow rate at the inlet. Some physiologic parameters are also given as stroke volume, heart rate, and cardiac output.

| $D_{STJ}$ [mm] | 26 | 30 | 34 |
| --- | --- | --- | --- |
| $\Delta x_{\min}^f$ [$\mu\text{m}$] | 87.4 | 75.6 | 75.6 |
| $\Delta x_{\max}^f$ [$\mu\text{m}$] | 765 | 804 | 828 |
| $\Delta x_{\text{avg}}^f$ [$\mu\text{m}$] | 458 | 495 | 503 |
| $Re$ | 6600 | 6600 | 6600 |
| $\eta^f$ [$\mu\text{m}$] | 32 | 32 | 32 |
| $\epsilon^f$ [$\text{cm}^2 \text{s}^{-3}$] | $3.3 \times 10^5$ | $3.3 \times 10^5$ | $3.3 \times 10^5$ |
| $Q_{\max}^f$ [$\text{cm}^3 \text{s}^{-1}$] | 405 | 405 | 405 |
| Systolic Volume [mL] | 83.33 | 83.33 | 83.33 |
| Heart rate [bpm] | 70 | 70 | 70 |
| Cardiac output [$\text{Lmin}^{-1}$] | 5.836 | 5.836 | 5.836 |

### 2.3 Flow Modeling

The blood was modeled as a Newtonian incompressible fluid with a density of *ρ*^*f*^ = 1.1 g cm^−3^ and a dynamic viscosity of *μ*^*f*^ = 3.5cPa. The Newtonian behavior approximation is valid in large blood vessels, where suspended particles (i.e., red blood cells) are far smaller than the characteristic sizes of the vessels (i.e., LVOT, aortic root, and aorta) [51]. Dirichlet boundary conditions were set at the inlet for the velocity, while at the outlet zero traction Neumann boundary conditions are imposed. The Navier-Stokes equations are discretized in space using the Finite Element (FE) method with linear elements and equal order interpolation for velocity and pressure, while an explicit 3^rd^-order Runge-Kutta scheme is used for the time discretization. Given that peak flow Reynolds numbers are around *Re* ∼ 6600, we consider a Large Eddy Simulation (LES) model to better capture the turbulent flow that characterizes the blood flow past TAVRs. In appendix D, the SA thrombogenic footprint is computed with and without an LES turbulence model for the flow. Differences were observed for the particles with the largest SAs, with higher stresses for the LES than for the laminar model, justifying the use of a turbulence model to avoid underestimating the stresses.

A Neohookean hyperelastic material model was used to model the TAVR leaflet material, with the Young modulus and Poisson ratio set to *E^s^* = 2MPa and *v^s^* = 0.3, respectively. The material parameters used by Oks et al. [50], which are consistent with the range of values employed by isotropic hyperelastic models of TAVR leaflets [52, 53]. The native aortic valves and the TAVR frame and cuff (see Figure 1b) were assumed rigid. The attachments of the leaflets to the stent were fixed, imposing zero Dirichlet boundary conditions on the displacement. As described in Oks et al. [50], a total Lagrangian formulation was used to implement the governing solid equations, which were discretized in space using the FE method and in time using an implicit generalized beta-Newmark scheme. Further details on the solid model, implementation, and chosen parameters can be found in [50].

Heart valves consist of soft tissue of density very similar to that of blood. Their mechanics are therefore dictated by an intricate non-linear feedback interaction between structure and flow. This makes it mandatory to consider 2-way FSI models to correctly reproduce the dynamics for the full cardiac cycle. This work used the immersed FSI coupling method introduced and validated in Oks et al. [50], which was designed to couple unstructured FE discretization of fluid and solid domains in problems involving large deformations of the fluid-solid interface. This method interpolates fluid velocities to the immersed solid and spreads out the solid internal forces to the fluid via a volumetric body-forcing term, to impose the no-slip boundary condition at the fluid-structure interface. A thorough description of this method can be found in [50].

The present FSI model was implemented in *Alya*, Barcelona Supercomputing Center’s in-house multi-physics simulation software designed to run efficiently on supercomputers. This software was verified, validated, and optimized to provide accurate solutions in complex fluid and solid mechanics problems, as well as other physics problems [54–60]. *Alya* has been developed to scale efficiently in parallel on CPUs and/or GPUs using hybrid MPI, OpenMP, CUDA, and/or OpenACC models. It is one of the twelve simulation codes of the Unified European Applications Benchmark Suite (UEABS) and thus complies with the highest standards in HPC [61]. A multi-code partitioned parallelization strategy was followed in this work, in which two instances of *Alya* were simultaneously executed, CFD in one instance and computational solid mechanics in another instance. The previous work by Oks et al. [50] discussed this strategy in further detail.

### 2.4 Lagrangian platelets tracking

To assess the risk of platelet activation by flow-induced stresses and their residence time in areas prone to stasis, platelets were modeled as Lagrangian particles seeded into the flow field near the inlet, with a random distribution and cutting off 4 mm away from the walls. The seeding was performed at regular intervals, 10 times per cardiac cycle, throughout 10 cardiac cycles. In each one of the 100 injections, 2×10^4^ Lagrangian particles were introduced into the domain, resulting in a total of 2×10^6^ particles introduced during the full simulation. A convergence analysis was carried out to assure that the number of particles injected was sufficiently large to assure the independence of the computed metrics from the seeded particles sample size. This analysis is presented in further detail in Appendix C. The platelets trajectories were solved using a one-way fluid-particle interaction coupling, considering drag and advection forces, in a Lagrangian frame of reference, i.e., following each platelet along its trajectory. Further details about platelets transport and deposition modeling are available in appendix A and literature [62, 63].

### 2.5 Hemodynamic characteristics

The orifice area is a measure of the degree of opening of the valve leaflets, illustrated in Figure 3a. It can be quantified using the Geometric Orifice Area (GOA), which is defined as the area enclosed by the projection of the leaflet commissure curves on the aortic cross-section (see Figure 3a). The GOA calculation was carried out at each stage of the cardiac cycle depicted in Figure 3c. The algorithm employed was further described in [50]. According to the ISO 5840-3 standard, Transvalvular Pressure Gradient (TPG) is an important indicator of prosthesis performance [38]. In this work, it was computed by averaging the fluid pressure at cross-section slices 2 cm upstream *P*_up_(*t*^*n*^) and 2 cm downstream *P*_down_ (*t*^*n*^) of the valve plane as shown in Figure 3b, and computing the difference between them at each time step *t*^*n*^: TPG (*t*^*n*^) = *P*_up_ (*t*^*n*^) − *P*_down_ (*t*^*n*^). The valve plane is the plane formed by the leaflet commissures in their initial state, that is, closed. As for the GOA, phase-averaged statistics are extracted from the TPG time series.

**Figure 3:**
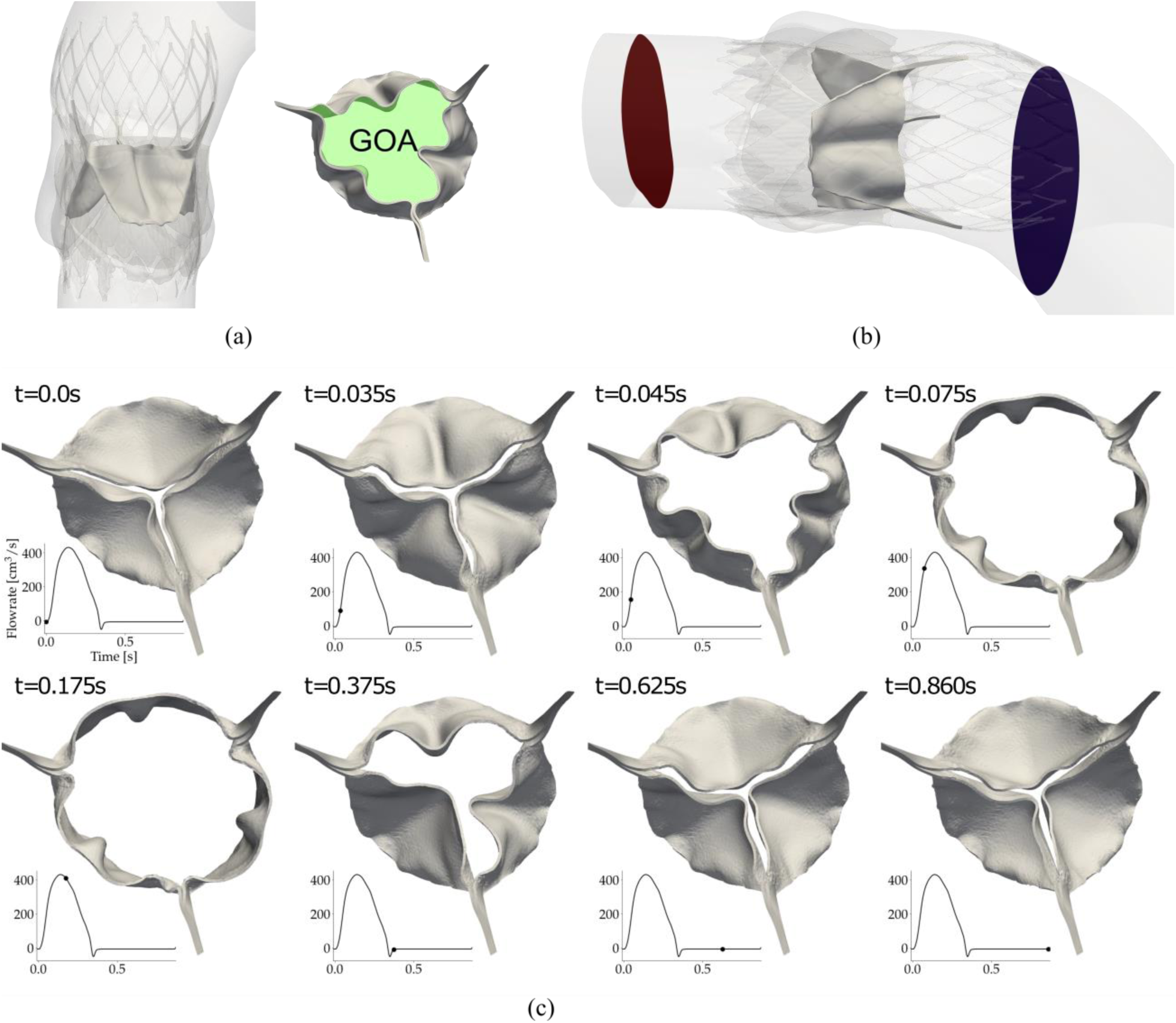
(a) GOA- the area enclosed by the projection of the leaflet edges on the valvular plane, shown in green, (b) Diagram of upstream (red) and downstream (blue) slices used to compute Transvalvular Pressure Gradient (TPG), (c) Leaflet motion during the cardiac cycle, with the dot indicating the time of the snapshot.

The Residence Time (*T_R_*) of a fluid tracer is the total time that the tracer has spent inside a control volume, quantifying the degree of washout of fluid at each point in space. Thrombosis is more likely to occur in low flow regions characterized by large *T_R_* [26, 64, 65]. In this work, *T_R_* was computed using a Eulerian approach: passive scalars were advected by the flow using equation (3) [66],

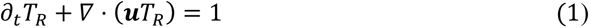

Dirichlet boundary conditions *T*_*R*_ = 0 were set at the inlet, while zero-flux Neumann boundary conditions, *n* ⋅ ∇*T*_*R*_ = 0, were set at the outlet and vessel walls (i.e., adiabatic boundary conditions). Details are described in literature [67]. Wall Shear Stress (WSS) was also measured at the aortic wall, and the surface of the stent.

To assess the stress history experienced by the platelets that act as an indicator of their activation state, the Stress Accumulation (SA) is calculated for each platelet along its trajectory [29]. By analyzing the distribution of stress accumulation value reached in each of these numerous platelet trajectories, a “thrombogenic footprint” of the TAVR device can be generated for each patient model. This “thrombogenic footprint” is represented by the Probability Density Function (PDF) of stresses accumulated on the platelets, as described in appendix B. Based on Hellums’ criterion, used in previous studies of thrombogenic risk, SA values > 3.5 Pa × s may drive the platelets beyond their activation threshold [30]. The platelets are here modeled as Lagrangian particles advected by the flow, considering drag forces, as explained in section 2.3. The stress tensor, defined as the symmetric gradient of the fluid velocities, is multiplied by the dynamic viscosity *μ*^*f*^:

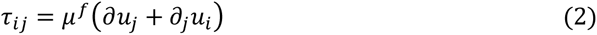

The components of the stress tensor *τ*_*ij*_ are reduced into a scalar stress, *σ*, based on the formulation of Bludszuweit [68] that is analogous to the von-Mises stress, typically used to predict the onset of yield in ductile solid materials.:

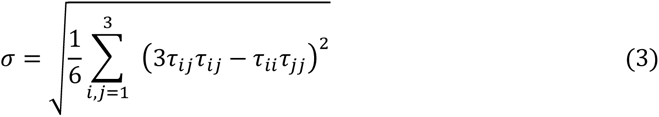

The scalar shear stress is then integrated over time along each particle trajectory 𝒞_traj._ . This integral is approximated by summing the instantaneous product of *σ* and the exposure time between timesteps Δ*t* of the platelet trajectory where *N*_time_ is the number of time steps.

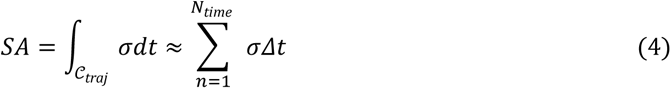

## Results

Results obtained from structural, hemodynamic, and thrombogenic risk analyses of the TAVR device in the three models with varying STJ are presented. A quantitative summary of several pertinent hemodynamic performance metrics obtained from the analysis is summarized in Table 2.

**Table 2:**
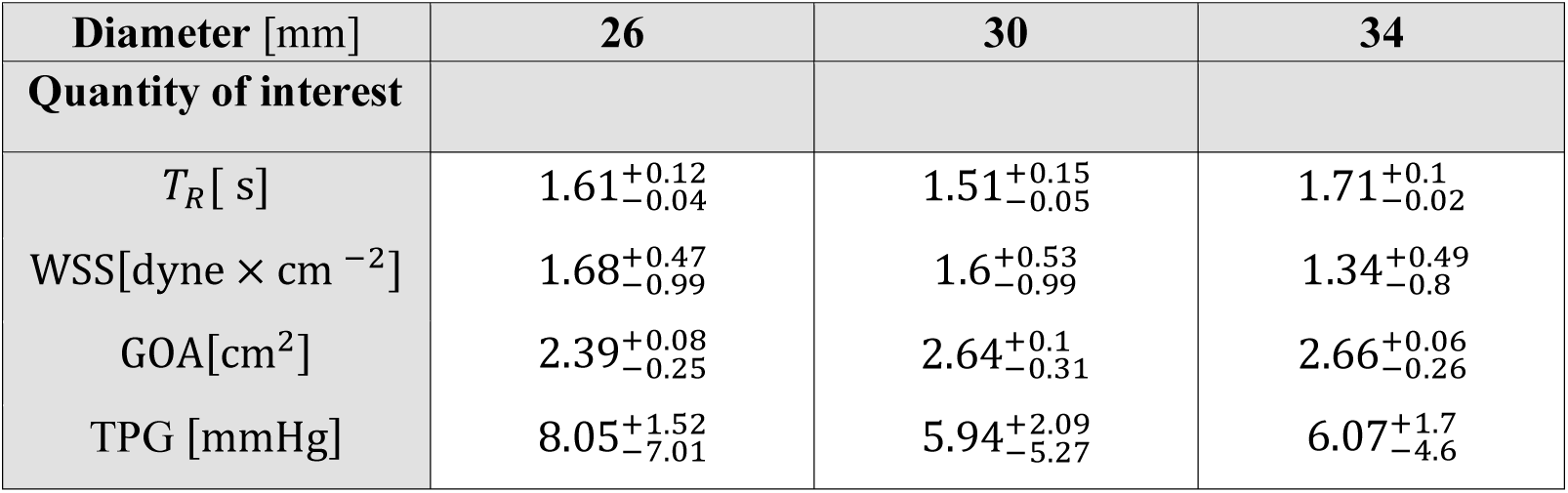
Results for time-statistics of volume-averaged quantities of interest. Values for Residence Time (T_R_), and Wall Shear Stress (WSS) correspond to the median computed over successive full cycles (shown with upper and lower deviations computed over the systolic periods of 2^nd^ to 10^th^ cardiac cycles).

| Diameter [mm] | 26 | 30 | 34 |
| --- | --- | --- | --- |
| Quantity of interest |  |  |  |
| $T_R$ [s] | $1.61^{+0.12}_{-0.04}$ | $1.51^{+0.15}_{-0.05}$ | $1.71^{+0.1}_{-0.02}$ |
| WSS[dyne × cm<sup>-2</sup>] | $1.68^{+0.47}_{-0.99}$ | $1.6^{+0.53}_{-0.99}$ | $1.34^{+0.49}_{-0.8}$ |
| GOA[cm<sup>2</sup>] | $2.39^{+0.08}_{-0.25}$ | $2.64^{+0.1}_{-0.31}$ | $2.66^{+0.06}_{-0.26}$ |
| TPG [mmHg] | $8.05^{+1.52}_{-7.01}$ | $5.94^{+2.09}_{-5.27}$ | $6.07^{+1.7}_{-4.6}$ |

### 2.6 TAVR implantation structural analysis

The expansion of the TAVR device was affected by the size of the STJ. In the crown region where the leaflet tips are connected, the cross-sectional area of the TAVR stents was reduced by 61%, 46%, and 33% for the models with STJ diameters of 26 mm, 30 mm, and 34 mm, respectively (as shown in Figure 4a). Additionally, Figure 4b illustrates that the cross-sectional area at the belly region of the TAVR stents is reduced by 13%, 8%, and 6% for the models with STJ diameters of 26 mm, 30 mm, and 34 mm, respectively. The reduction in cross-sectional area at the left ventricular outflow tract region was similar for all three models, at approximately 40% (Figure 4c).

**Figure 4:**
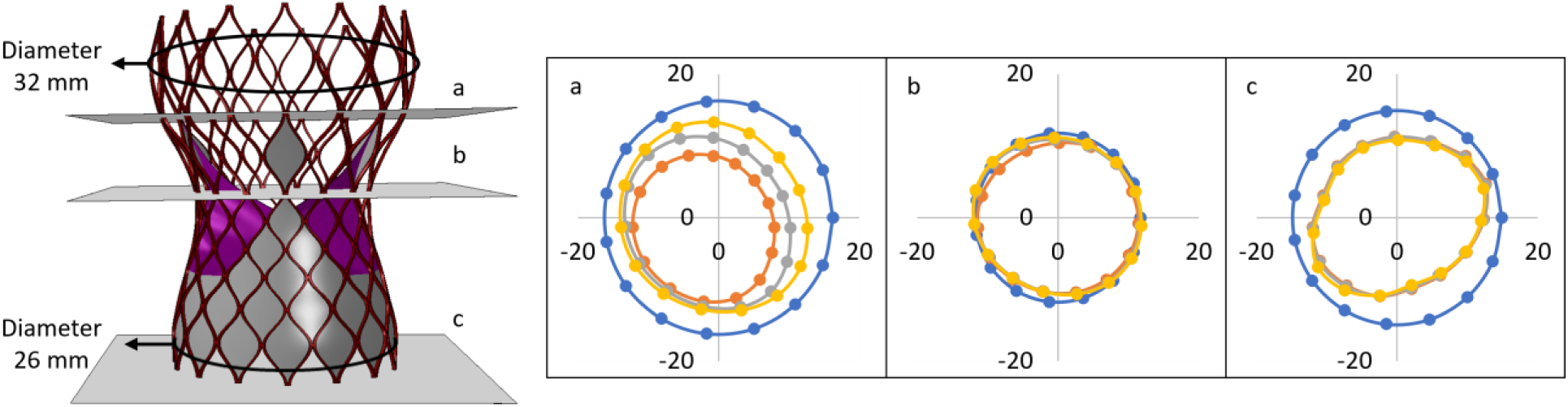
Cross-sections of the undeformed (blue), and deformed stents implanted inside models with 26 mm (red), 30 mm (grey), and 34 mm (yellow) STJ at the joint of leaflet tip and crown of the stent (a), the belly of the stent (b), and bottom end of the stent (c). The cross-sections are presented in mm scale.

### 2.7 Device hemodynamic performance

The pathological anatomical features found in CAVD patients lead to complex flow patterns are depicted in Figure 5. Performance of prosthetic heart valves typically involves analyzing various parameters, such as the systolic orifice area and the TPG, as specified in ISO 5840-3 [38]. Figure 5a displays the velocity distribution, while Figure 5b illustrates the pressure distribution during peak systole for the three models with varying STJ sizes. The effect of the STJ size manifests in the different jet formation dynamics and flow patterns formed downstream. In the model with the smallest STJ, a high-velocity jet was observed, which propels the major vortex away from the region during systole. As the STJ size increased, the major vortex was observed to transport more slowly through the aortic outflow tract, increasing the flow stagnation duration inside the Sinus of Valsalva (SoV). (Supplementary animations 1 and 2).

**Figure 5:**
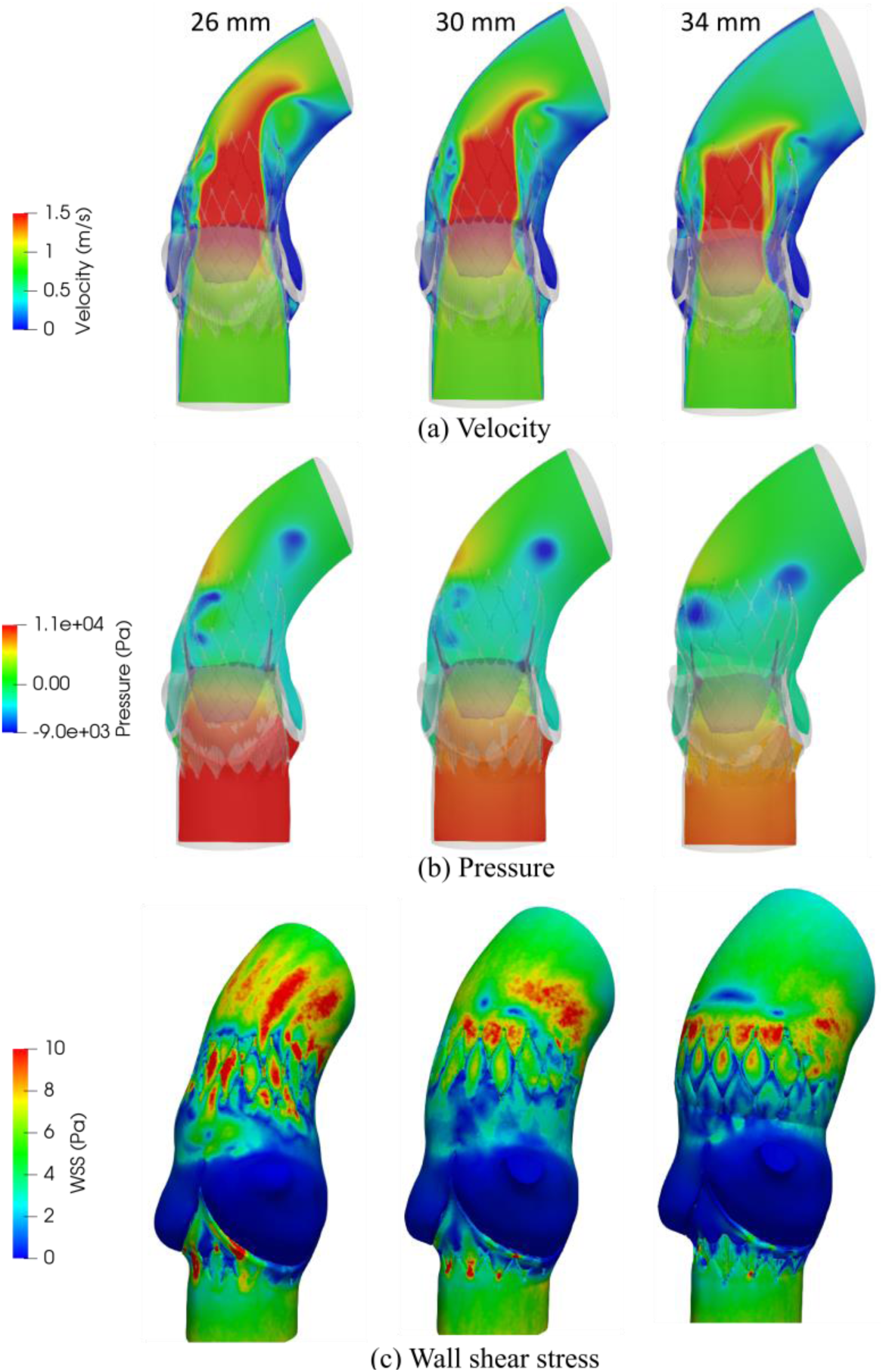
Snapshots of (a) velocity, (b) pressure, (c) wall shear stress contours, for 26, 30, and 34 mm STJ diameter parametric models during systole.

The Geometric Orifice Area (GOA) was computed for each model with different STJ diameters as described in section 2.5. The orifice area of the 30 mm and 34 mm STJ models were comparable (Table 2). However, the GOA was decreased for the smallest STJ, approximately 10 % as compared to the larger STJ 30 mm and 34 mm diameters models. The larger outflow produced a lower downstream pressure, facilitating the easier opening of the leaflets. The systolic Transvalvular Pressure Gradient (TPG) decreased for larger STJ models, with a reduction of 25% from the smallest to the largest STJ models as indicated in Table 2. Higher TPGs typically lead to reduced ejection fractions and higher pressure loads on the left ventricle. Therefore, the ISO 5840-3 standard [38] defines critical maximum thresholds for devices.

Snapshots for WSS fields are shown in Figure 5c, that depicts a decrease in WSSs for larger STJs. For the narrowest geometry (26 mm STJ model), stresses are highly concentrated on the stents which can be seen to produce the onset of WSS streaks at the aortic outflow tract. Moreover, Table 2 shows a 20% decrease of WSS for 34 mm STJ diameter model compared to 26 mm STJ diameter model.

### 2.8 Device thrombus formation and thrombogenic risk analysis

The Residence Time (*T_R_*) is an indicator of the degree of flow stasis that may promote thrombus formation. Figure 6a shows vertical and horizontal cross-sections of the *T_R_* distribution patterns for the three models during early systole of the 10^th^ cardiac cycle. The longest *T_R_*s are observed in the cusps of the sinus of Valsalva and Neosinus. Figure 6a shows that the right and left coronary cusps of the 26 mm STJ model had relatively higher local *T_R_* compared to the 30 mm STJ model due to a partial obstruction of washout caused by the native valve leaflets which are pushed against the walls by the TAVR stent. However, a strong jet close to the non-coronary cusp enabled an efficient washout at the non-coronary cusp of the 26 mm STJ model compared to 30 mm and 34 mm STJ models. An overall increase in *T_R_* can be observed in the SoV and Neosinus regions (Figure 6a) for the 34 mm STJ model compared to 30 mm STJ model which indicates that the model with 30 mm STJ experienced the shortest *T_R_*. Comparing the model with a 34 mm STJ diameter to the one with a 26 mm STJ diameter reveals an increase of 6% in the median of volume-averaged *T_R_* (Table 2)

**Figure 6:**
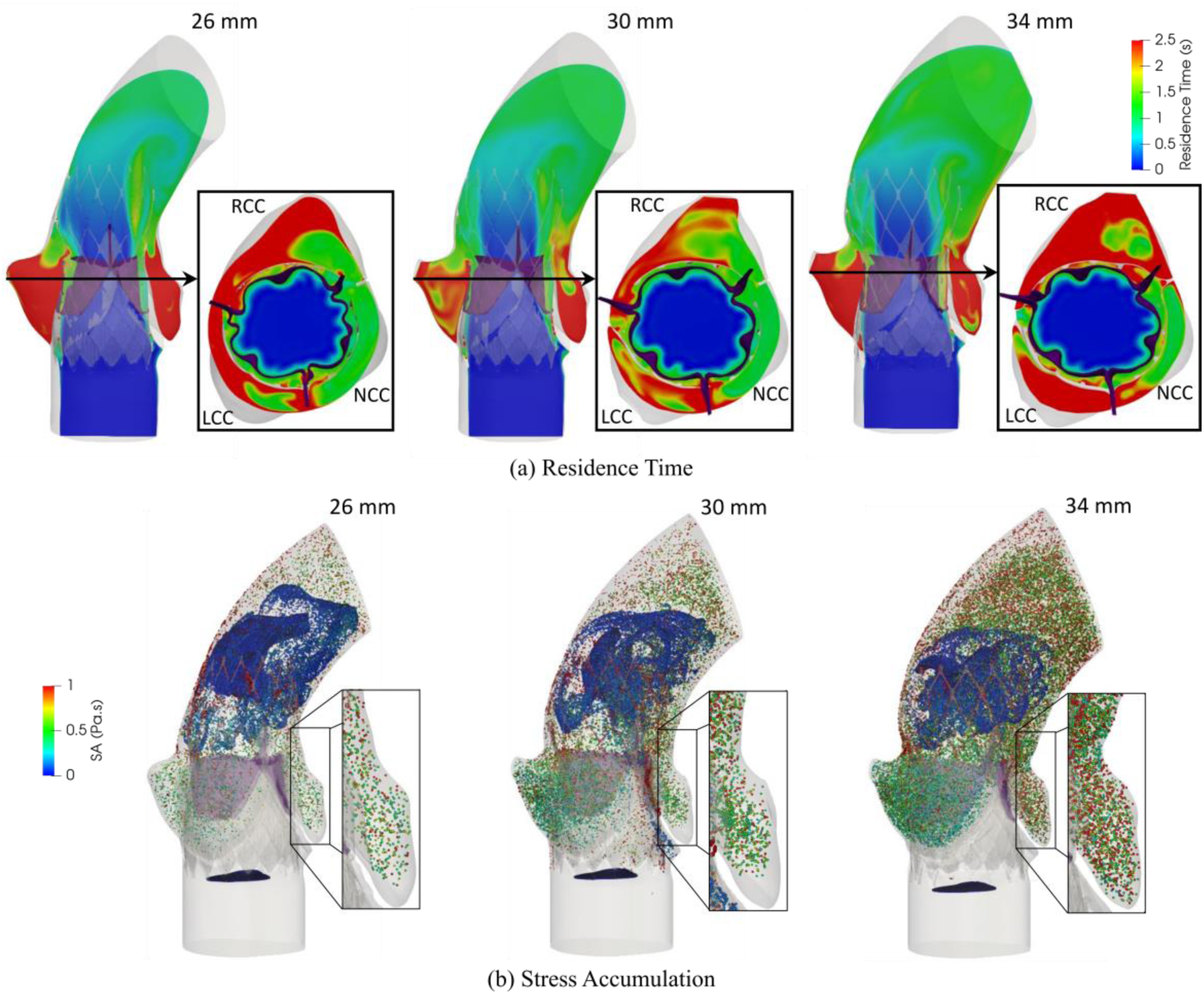
(a) Axial and cross-sectional snapshots of residence time (b) stress accumulation of particles seeded near the inlet of the domain, with a diameter of 3µm. Zoom-ins show the number of platelets with high SA in the sinus region, for 26, 30, and 34 mm STJ diameter models during early systole of the 10^th^ cardiac cycle. NCC= Non coronary cusp, LCC= Left coronary cusp, and RCC= Right coronary cusp. (Particles are scaled up for visualization).

Stress Accumulation analysis in Lagrangian framework indicated that the incidence of platelets exhibiting higher SA values in the SoV and Neosinus regions is more prominent in models with larger STJ sizes (Figure 6b). In order to compare this trend quantitatively, we conducted a comparative analysis among the three models with varying STJ sizes, the SA values for all platelets for each model were collapsed into a probability density function (PDF). Figure 7 shows the PDFs for the stress accumulations of platelets in each one of the regions of interest for analyzing the thrombogenic risk: the Sinus of Valsalva and the Neo sinus. Zoomed-in insets are provided for the SA tail range of the PDF distribution that exceeds 10^−1^ to 10 Pa×s - representing the most thrombogenic platelets in the ensemble. In both regions of interest, a majority of platelets have SA values below 10^−2^ Pa×s, indicating a low risk of platelet activation. The portion of platelets within this range increases with the STJ diameter. Conversely, for intermediate SA values ranging from 10^−2^ to 10^−1^ Pa×s, this trend is reversed, and the portion of platelets decreases with larger STJ diameters. The trend is once again reversed at the tail of the PDF which is the range associated with the highest potential for platelet activation, i.e., above 10^−1^ Pa×s. The portion of platelets in this range increases with larger STJ diameters mainly due to an elevated residence time within the domain (refer to insets in Figure 7).

**Figure 7:**
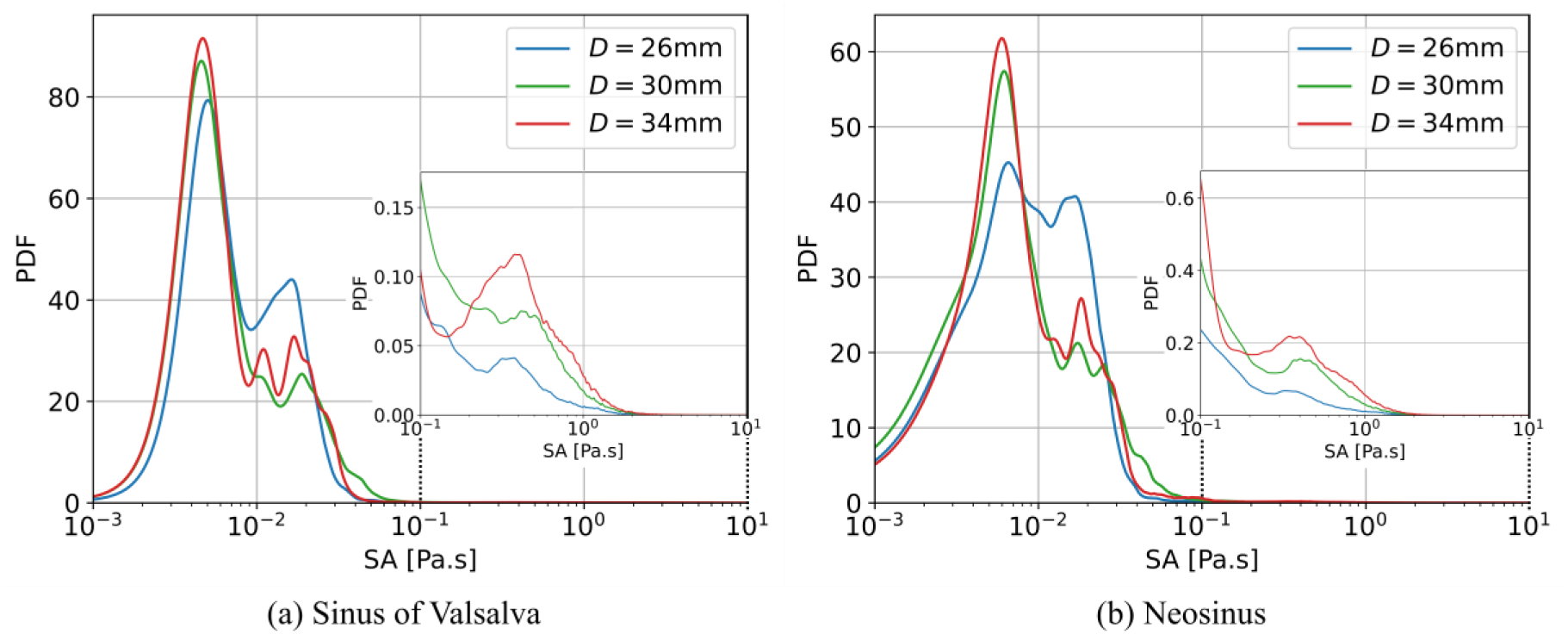
Probability density function of stress accumulation distributions for 26 mm (blue), 30 mm (green), and 34 mm (red) STJ diameter geometries computed in (a) sinus of Valsalva, and (b) neosinus. These distributions can be interpreted as the thrombogenic footprint of each device-patient configuration. The horizontal axis is in logarithmic scale for clarity.

## 3 Discussion

This study analyzed the structural and hemodynamic impact of STJ diameter in TAVR devices implanted supra-annularly. The structural analysis indicated that the smallest STJ size (26 mm) reduced the cross-sectional area of the TAVR at the crown region by more than a half. Reduction in cross-sectional area depicts the under-expansion of the stent struts in leaflet tip and belly regions that may affect the leaflet dynamics. The hemodynamic analysis confirms this by illustrating that the smaller STJ diameter impairs the hemodynamic performance of supra-annular TAVR devices by reducing the available GOA and elevating the TPG. This smaller GOA generates a narrow high-velocity jet that impinges the endothelial lining at the ascending aorta and elevating the WSS. Elevated WSS at the ascending aorta often leads to endothelial damage and further complications. The diameter of the considered stent at the crown region was 32 mm which is structurally favorable for the 30 mm STJ model with an STJ cover index closest to 1 (0.94). The 34 mm STJ on the other hand allows the stent to have a moderately larger opening at the crown region that translates into a marginal increase in GOA, resulting in a relatively lower jet velocity and WSS as compared to the 30 mm STJ.

The incidence of thromboembolic events in TAVR devices primarily occurs due to abnormal blood flow patterns through narrowed conduits leading to increased shear stresses on platelets and prolonged exposure time in recirculation zones where platelets get trapped. We analyzed flow stasis and stagnation in a Eulerian framework and additionally applied the DTE approach, which considers the combined effect of the shear stress and the duration of platelet exposure to it, i.e., the residence time. In the case of the 26 mm STJ model, an incomplete vortex formation during systole was observed, which propelled the majority of platelets out of the Sinus of Valsalva (SoV) and beyond the domain. Conversely, larger STJ models exhibited a prominent vortex that moved relatively slower along the domain, leading to increased flow stagnation in both the SoV and the Neosinus as the STJ size increased. Consistently, a larger number of platelets with higher SA magnitudes were observed within the SoV and Neosinus regions of the larger STJ models, as manifested in the zoom-ins of Figure 6b. Interestingly, the 26 mm STJ model demonstrated high local stagnation zones at the LCC and RCC due to narrower sinus openings caused by the TAVR prosthesis pushing the native leaflets closer to the aortic wall. This can potentially lead to coronary occlusion, increasing the risk of myocardial infarction [69]. These observations are analyzed by the stress accumulation on advected platelets, as depicted in Figure 7.

The statistical distribution (PDF) of seeded platelets’ stress accumulation (SA) depends both on the patient-device scenarios and the different STJ diameters. In both RoIs (Sinus of Valsalva and Neosinus), most platelets reside in the lower SA range with a low risk of platelet activation. The portion of platelets in this range increases with increasing STJ diameter. For intermediate SA range, this tendency is inverted, and the portion of platelets at this SA range decreased with the STJ diameter. Smaller STJ diameters generate stronger jets; however, those induce recirculation formation further downstream in the aortic outflow tract, yielding more platelets with SA values within this intermediate range, resulting in the number of platelets at this range inversely correlated with the STJ diameter. These platelets may experience further free flow shear stress downstream and may reach the critical activation threshold that may result in thromboembolism. Finally, this trend is reversed once again with the portion of platelets at the tail of the PDF, associated with elevated platelet-activation potential. Larger STJ diameters provide larger space for recirculation formations trapping a large number of the platelets inside the domain that accumulate stress because of the increased residence time (Figure 6b). Platelets in this range are more likely to be activated inside the domain and may cause HALT. It is worth mentioning that other factors such as paravalvular leakage, unique patient-specific anatomical features, or biochemistry may further increase the thrombogenic risk [34]. No significant fraction of the PDF distribution is above the critical platelet activation threshold of 3.5 Pa×s. However, this threshold that was established by Hellums for various combinations of constant shear stress and exposure times is not necessarily translatable to all dynamic conditions and scenarios. Moreover, the PDF thrombogenic footprint is most useful for comparative applications, contrasting the different distributions to further elucidate the effect of different patient-specific anatomies and devices on the stress experienced by platelets in blood flow [16].

In clinical practice, self-expandable devices are oversized with respect to the annulus diameter to ensure proper anchorage and minimize paravalvular leakage. Patients with smaller STJ size are generally recommended for relatively smaller device oversizing. However, clinical studies have indicated that patients with smaller STJ cover index (STJCI) are prone to device malposition, and paravalvular leakage [42]. In this study, we observed that the STJ size affects the structural implantation, hemodynamic performance, and thrombogenic risk of supra-annular TAVR devices. In summary, the 26 mm Evolut^®^ supra-annular TAVR device performed the best for the patient with STJCI closest to 1 (30 mm STJ size) with an optimal GOA, TPG, residence time, and thrombogenic footprint. Narrow STJ (STJCI<<1) caused under-expansion of the TAVR that led to inferior hemodynamic performance with increased risk of coronary occlusion indicating potential thromboembolic complications downstream. Patients with a narrower range of STJ size may require consideration for alternative procedures. In contrast, larger STJ (STJCI>1) facilitated larger expansion of the TAVR prosthesis at the crown region and demonstrates comparable hemodynamic performance to the patient with 30 mm STJ. However, larger STJs produce larger stasis regions that allows more platelets already with high SA values to remain in the Neosinus that may lead to HALT, and/or leaflet degeneration. The patients with larger STJ size may need more aggressive anticoagulation therapy.

The present study provides a novel refined computing framework that evaluates the impact of patient-specific anatomic parameters on the thrombogenic risk associated with TAVR and the potential for HALT formation. Moreover, the study highlights the value of employing virtual patient populations to elucidate the influence of anatomical parameters on device performance and associated risks. Specifically, the development of subclinical leaflet thrombosis poses significant concerns for patients, clinicians, and device manufacturers, and addressing it is of utmost importance. The introduced framework holds significant potential in addressing this critical need, with the goal of improving preprocedural planning of TAVR devices for better clinical outcomes, ultimately saving lives.

## 4 Limitations

Although this study presents noteworthy contributions, it is important to recognize some limitations. One patient model was considered for this study along with two virtual patient cohorts with varying STJ sizes to study the effect of the STJ size, while keeping the other anatomical and procedural parameters unchanged. While numerous studies have emphasized the need of using virtual patient cohorts in boosting the capacity to conduct diagnosis, prognosis, and procedural planning [43, 70], the application of our methodology to a specific TAVR device is a potential limitation. A large patient cohort may be required to confirm the findings presented. Nevertheless, this study serves as a methodological approach that combines highly complex and computationally demanding methods (patient-specific model reconstruction, FEA, FSI, Lagrangian particle tracking, etc.) to assess the device thrombogenic risk associated with an anatomical parameter (STJ size). The presented framework can be used to study the effect of other anatomical and procedural parameters on the thrombogenic risk of TAVR.

The FEA-based TAVR deployment simulation assumed a uniform aortic wall thickness, whereas in wall thickness may vary. The aortic wall, native leaflets, as well as the TAVR device porcine pericardium leaflets were simulated using isotropic hyperelastic material models, assuming that the anisotropic behavior of the leaflets is negligible. This assumption is supported by our previous studies where we demonstrated that these material models effectively approximate TAVR deployment simulations, exhibiting close agreement with *in vitro* results [48]. In the FSI simulations rigid aortic walls and stents were assumed, assuming that their compliance is negligible during the cardiac cycle. In the CFD model used for studying the thrombogenic risk, pressure in the ascending aorta changes dynamically due to the compliance and resistance offered by the systemic circulation. However, given the complexity of the simulations involved we applied a set constant pressure boundary condition at the outlets of the domain (and a flow rate waveform at the inlet). While this assumption is commonly used in such simulations, it may affect the values of TPGs.

## 5 Conclusions

In this study, patient-specific TAVR device deployment, hemodynamic performance, thrombogenic risk and the potential for thrombus formation that may lead to subclinical leaflet thrombosis leading to Hypoattenuated Leaflet Thickening (HALT) were simulated in a supra-annular Medtronic Evolut^®^ TAVR device, in order to examine the relation between the Sinotubular Junction (STJ) diameter used by clinicians for TAVR sizing, to the resultant device performance. The abovementioned metrics were rigorously analyzed to assess their effects on the potential clinical outcomes of the procedure in respect to the STJ sizing criteria. Structural analysis of the device deployment, fluid-structure interaction of the hemodynamic performance and CFD analysis of the thrombogenic potential were performed in three virtual patient cohorts of a patient-specific model with varying diameters of the STJ. The studies revealed an intricate interplay between the anatomical and procedural parameters that affects the hemodynamic performance and the risk of leaflet thrombosis. The model with STJCI closest to 1 (30 mm STJ model) exhibited optimal structural and hemodynamic performance. Under-expansion of the TAVR device in the smallest STJ model resulted in inferior hemodynamic performance with a potential risk of thromboembolic complications downstream. In contrast, the 34 mm STJ model demonstrated comparable hemodynamic performance to the best performing 30 mm STJ model, with the exception of higher count of platelet with stress accumulation captured in the neo-sinuses, indicating an elevated risk of leaflet thrombosis that may lead to HALT. This refined biomechanical analysis may aid clinicians in enhancing their pre-procedural planning for TAVR and making informed decisions regarding post-procedural anticoagulant therapy, while considering each patient’s unique anatomical characteristics.

## Acronyms

TR: Residence Time.
AVR: Aortic Valve Replacement.
CFD: Computational Fluid Dynamics.
CT: Computed Tomography.
DTE: Device Thrombogenicity Emulation.
FE: Finite Element.
FEA: Finite Element Analysis.
FSI: Fluid-Structure Interaction.
GOA: Geometric Orifice Area.
HALT: Hypoattenuated Leaflet Thickening.
LES: Large Eddy Simulation.
LVOT: Left ventricle outflow tract.
PDF: Probability Density Function.
RoI: Region of Interest.
SA: Stress Accumulation.
SAVR: Surgical Aortic Valve Replacement.
SoV: Sinus of Valsalva.
STJ: Sinotubular Junction.
TAVR: Transcatheter Aortic Valve Replacement.
TPG: Transvalvular Pressure Gradient.
WSS: Wall Shear Stress.

## Author Contributions

S.R. and D.O. conducted the study and prepared the manuscript. G.H. and C.S. contributed significantly to the design and implementation of algorithms. C.S., M.V., B.K., and D.B. contributed to the critical review, interpretation of results, and the writing.

## Ethics Declarations

The study was conducted in accordance with the Declaration of Helsinki and approved by the Institutional Review Board of Stony Brook University (2013-2357-R5, 2 October 2022).

## Data availability

Data is contained within the article and Appendix.

## References

1. iData, Cardiac Surgery Market Size, Share, and COVID-19 Impact Analysis | United States | 2020-2026 | MedSuite | Includes: Tissue Heart Valve Market, TAVI/TAVR Market, and 21 more. 2022.

2. Yudi, M.B., et al., Coronary angiography and percutaneous coronary intervention after transcatheter aortic valve replacement. Journal of the American College of Cardiology, 2018. 71(12): p. 1360–1378.

3. Adams, D.H., et al., Transcatheter aortic-valve replacement with a self-expanding prosthesis. New England Journal of Medicine, 2014. 370(19): p. 1790–1798.

4. Leon, M.B., et al., Transcatheter or surgical aortic-valve replacement in intermediate-risk patients. New England Journal of Medicine, 2016. 374(17): p. 1609–1620.

5. Popma, J.J., et al., Transcatheter aortic valve replacement using a self-expanding bioprosthesis in patients with severe aortic stenosis at extreme risk for surgery. Journal of the American College of Cardiology, 2014. 63(19): p. 1972–1981.

6. Nishimura, R.A., et al., 2014 AHA/ACC Guideline for the Management of Patients With Valvular Heart Disease: Executive Summary. Circulation, 2014. 129(23): p. 2440–2492.

7. Vahanian, A., et al., ESC Committee for Practice Guidelines (CPG); Joint Task Force on the Management of Valvular Heart Disease of the European Society of Cardiology (ESC); European Association for Cardio- Thoracic Surgery (EACTS). Eur J Cardiothorac Surg., 2012. 42(4): p. S1–44.

8. Chakravarty, T., et al., Subclinical leaflet thrombosis in surgical and transcatheter bioprosthetic aortic valves: an observational study. The Lancet, 2017. 389: p. 2383–2392.

9. Jose, J., et al., Clinical Bioprosthetic Heart Valve Thrombosis After Transcatheter Aortic Valve Replacement: Incidence, Characteristics, and Treatment Outcomes. JACC: Cardiovascular Interventions, 2017. 10(7): p. 686–697.

10. Mangione, F.M., et al., Leaflet Thrombosis in Surgically Explanted or Post-Mortem TAVR Valves. JACC: Cardiovascular Imaging, 2017. 10(1): p. 82–85.

11. Makkar, R.R., et al., Subclinical Leaflet Thrombosis in Transcatheter and Surgical Bioprosthetic Valves. Journal of the American College of Cardiology, 2020. 75(24): p. 3003–3015.

12. Makkar, R.R., et al., Possible Subclinical Leaflet Thrombosis in Bioprosthetic Aortic Valves. New England Journal of Medicine, 2015. 0(0): p. null.

13. Nagpal, P., et al., Imaging of the aortic root on high-pitch non-gated and ECG-gated CT: awareness is the key! Insights into Imaging, 2020. 11.

14. Nappi, F., et al., Are the dynamic changes of the aortic root determinant for thrombosis or leaflet degeneration after transcatheter aortic valve replacement? Journal of Thoracic Disease, 2020. 12(5).

15. Hsiung, I., et al., Left Main Protection During Transcatheter Aortic Valve Replacement With a Balloon-Expandable Valve. Journal of the Society for Cardiovascular Angiography & Interventions, 2022: p. 100339.

16. Pan, Y., A. Qiao, and N. Dong, Fluid–Structure Interaction Simulation of Aortic Valve Closure with Various Sinotubular Junction and Sinus Diameters. Annals of Biomedical Engineering, 2014. 43: p. 1363–1369.

17. Marom, G., et al., Numerical model of the aortic root and valve: Optimization of graft size and sinotubular junction to annulus ratio. The Journal of Thoracic and Cardiovascular Surgery, 2013. 146(5): p. 1227–1231.

18. Casa, L.D.C., D.H. Deaton, and D.N. Ku, Role of high shear rate in thrombosis. Journal of Vascular Surgery, 2015. 61(4): p. 1068–1080.

19. Hellums, J.D., 1993 Whitaker lecture: Biorheology in thrombosis research. Annals of Biomedical Engineering, 2006. 22: p. 445–455.

20. Casa, L.D.C. and D.N. Ku, Thrombus Formation at High Shear Rates. Annual Review of Biomedical Engineering, 2017. 19(1): p. 415–433.

21. Ducci, A., et al., Transcatheter aortic valves produce unphysiological flows which may contribute to thromboembolic events: An in-vitro study. Journal of Biomechanics, 2016. 49: p. 4080–4089.

22. Ghosh, R.P., et al., Numerical evaluation of transcatheter aortic valve performance during heart beating and its post-deployment fluid--structure interaction analysis. Biomechanics and modeling in mechanobiology, 2020. 19(5): p. 1725–1740.

23. Hatoum, H., et al., Aortic sinus flow stasis likely in valve-in-valve transcatheter aortic valve implantation. The Journal of Thoracic and Cardiovascular Surgery, 2017. 154: p. 32–43.

24. Midha, P.A., et al., The Fluid Mechanics of Transcatheter Heart Valve Leaflet Thrombosis in the Neosinus. Circulation, 2017. 136(17): p. 1598–1609.

25. Vahidkhah, K. and A.N. Azadani, Supra-annular Valve-in-Valve implantation reduces blood stasis on the transcatheter aortic valve leaflets. Journal of Biomechanics, 2017. 58: p. 114–122.

26. Vahidkhah, K., et al., Valve thrombosis following transcatheter aortic valve replacement: significance of blood stasis on the leaflets. Eur J Cardiothorac Surg, 2017. 51(5): p. 927–935.

27. Bluestein, D., S. Einav, and M.J. Slepian, Device thrombogenicity emulation: A novel methodology for optimizing the thromboresistance of cardiovascular devices. J Biomech, 2012.

28. Girdhar, G., et al., Device thrombogenicity emulation: a novel method for optimizing mechanical circulatory support device thromboresistance. PLoS One, 2012: p. e32463.

29. Alemu, Y. and D. Bluestein, Flow-induced platelet activation and damage accumulation in a mechanical heart valve: Numerical studies. Artificial Organs, 2007. 31(9): p. 677–688.

30. Xenos, M., et al., Device Thrombogenicity Emulator (DTE) - Design optimization methodology for cardiovascular devices: A study in two bileaflet MHV designs. Journal of Biomechanics, 2010. 43(12): p. 2400–2409.

31. Yin, W., et al., Flow-induced platelet activation in bileaflet and monoleaflet mechanical heart valves. Ann Biomed Eng, 2004. 32: p. 1058–1066.

32. Marom, G., et al., Numerical Model of Full-Cardiac Cycle Hemodynamics in a Total Artificial Heart and the Effect of Its Size on Platelet Activation. J. Cardiovasc. Transl. Res, 2014. 7: p. 788–796.

33. Anam, S.B., et al., Assessment of paravalvular leak severity and thrombogenic potential in transcatheter bicuspid aortic valve replacements using patient-specific computational modeling. Journal of Cardiovascular Translational Research, 2021: p. 1–11.

34. Bianchi, M., et al., Patient-specific simulation of transcatheter aortic valve replacement: impact of deployment options on paravalvular leakage. Biomechanics and modeling in mechanobiology, 2019. 18(2): p. 435–451.

35. Kovarovic, B.J., et al., Mild Paravalvular Leak May Pose an Increased Thrombogenic Risk in Transcatheter Aortic Valve Replacement (TAVR) Patients-Insights from Patient Specific In Vitro and In Silico Studies. Bioengineering, 2023. 10(2): p. 188.

36. Hatoum, H., et al., Predictive model for thrombus formation after transcatheter valve replacement. Cardiovascular Engineering and Technology, 2021. 12(6): p. 576–588.

37. Singh-Gryzbon, S., et al., Influence of patient-specific characteristics on transcatheter heart valve neo- sinus flow: an in silico study. Annals of biomedical engineering, 2020. 48: p. 2400–2411.

38. for Standardization, I.O., Cardiovascular Implants – Cardiac Valve Prostheses – Part 3: Heart Valve Substitutes Implanted by Transcatheter Techniques - ISO 5840-3:2013(E). 2013, International Organization for Standardization: Geneva, CH.

39. Reza, S., et al., A computational framework for post-TAVR cardiac conduction abnormality (CCA) risk assessment in patient-specific anatomy. Artificial Organs, 2022. 46(7): p. 1305–1317.

40. Yushkevich, P., et al., Yushkevich PA, Piven J, Hazlett HC, Smith RG, Ho S, Gee JC, Gerig GUser-guided 3D active contour segmentation of anatomical structures: significantly improved efficiency and reliability. Neuroimage 31:1116-1128. NeuroImage, 2006. 31: p. 1116–28.

41. Smith, M., ABAQUS/Standard User’s Manual, Version 6.9. 2009, United States: Dassault Systèmes Simulia Corp.

42. Wang, Y., et al., Anatomic predictor of severe prosthesis malposition following transcatheter aortic valve replacement with self-expandable Venus-A Valve among pure aortic regurgitation: A multicenter retrospective study. Frontiers in Cardiovascular Medicine, 2022. 9: p. 1002071.

43. Romero, P., et al., Clinically-driven virtual patient cohorts generation: An application to aorta. Frontiers in Physiology, 2021: p. 1375.

44. Hamdan, A., et al., Sex differences in aortic root and vascular anatomy in patients undergoing transcatheter aortic valve implantation: a computed-tomographic study. Journal of cardiovascular computed tomography, 2017. 11(2): p. 87–96.

45. Morganti, S., et al., Prediction of patient-specific post-operative outcomes of TAVI procedure: The impact of the positioning strategy on valve performance. Journal of Biomechanics, 2016.

46. Martin, C., T. Pham, and W. Sun, Significant differences in the material properties between aged human and porcine aortic tissues. European Journal of Cardio-Thoracic Surgery, 2011. 40(1): p. 28–34.

47. Martin, C. and W. Sun, Biomechanical characterization of aortic valve tissue in humans and common animal models. Journal of biomedical materials research Part A, 2012. 100(6): p. 1591–1599.

48. Anam, S.B., et al., Validating in silico and in vitro patient-specific structural and flow models with transcatheter bicuspid aortic valve replacement procedure. Cardiovascular Engineering and Technology, 2022: p. 1–17.

49. Systems, B.C., ANSA pre-processor. 2021.

50. Oks, D., et al., Fluid-structure interaction analysis of eccentricity and leaflet rigidity on thrombosis biomarkers in bioprosthetic aortic valve replacements. bioRxiv, 2022.

51. Sochi, T., Non-Newtonian Rheology in Blood Circulation. arXiv: Fluid Dynamics, 2013.

52. Bavo, A.M., et al., Fluid-Structure Interaction Simulation of Prosthetic Aortic Valves: Comparison between Immersed Boundary and Arbitrary Lagrangian-Eulerian Techniques for the Mesh Representation. PLoS ONE, 2016. 11(4): p. e0154517.

53. Kandail, H.S., et al., Impact of annular and supra-annular CoreValve deployment locations on aortic and coronary artery hemodynamics. J Mech Behav Biomed Mater, 2018. 86: p. 131–142.

54. Lehmkuhl, O., et al., A low-dissipation finite element scheme for scale resolving simulations of turbulent flows. Journal of Computational Physics, 2019. 390: p. 51–65.

55. Calmet, H., et al., Large-scale CFD simulations of the transitional and turbulent regime for the large human airways during rapid inhalation. Computers in Biology and Medicine, 2016. 69: p. 166–180.

56. Mira, D., et al., Heat Transfer Effects on a Fully Premixed Methane Impinging Flame. Flow Turbulence Combust, 2016. 97: p. 339–36.

57. Vázquez, M., et al., Alya: Multiphysics engineering simulation toward exascale. Journal of Computational Science, 2016. 14: p. 15–27.

58. Gövert, S., et al., Heat loss prediction of a confined premixed jet flame using a conjugate heat transfer approach. International Journal of Heat and Mass Transfer, 2017. 107: p. 882–894.

59. Santiago, A., et al., Design and execution of a Verification, Validation, and Uncertainty Quantification plan for a numerical model of left ventricular flow after LVAD implantation. PLoS Comput Biol., 2021.

60. Rodriguez, I., et al., Fluid dynamics and heat transfer in the wake of a sphere. International Journal of Heat and Fluid Flow, 2019. 76: p. 141–153.

61. Oyarzun, G., D. Mira, and G. Houzeaux, Performance assessment of CUDA and OpenACC in large scale combustion simulations. arXiv [cs.DC], 2021.

62. Calmet, H., et al., Flow features and micro-particle deposition in a human respiratory system during sniffing. Journal of Aerosol Science, 2018. 123: p. 171–184.

63. Calmet, H., et al., Subject-variability effects on micron particle deposition in human nasal cavities. Journal of Aerosol Science, 2018. 115: p. 12–28.

64. Rayz, V.L., et al., Flow residence time and regions of intraluminal thrombus deposition in intracranial aneurysms. Ann Biomed Eng., 2010. 38(10): p. 3058–3069.

65. Maud, B.G. and V.S. Michael, Biomaterial-associated thrombosis: roles of coagulation factors, complement, platelets and leukocytes. Biomaterials, 2004. 25(26): p. 5681–5703.

66. Reza, M.M.S. and A. Arzani, A critical comparison of different residence time measures in aneurysms. J Biomech, 2019. 88: p. 122–129.

67. Rossini, L., et al., A clinical method for mapping and quantifying blood stasis in the left ventricle. Journal of Biomechanics, 2016. 49(11): p. 2152–2161.

68. Bludszuweit, C., Model for a general mechanical blood damage prediction. Artificial Organs, 1995. 19: p. 583–9.

69. Thygesen, K., et al., Fourth Universal Definition of Myocardial Infarction (2018). Circulation, 2018. 138(20): p. e618–e651.

70. Niederer, S.A., et al., Creation and application of virtual patient cohorts of heart models. Philosophical Transactions of the Royal Society A, 2020. 378(2173): p. 20190558.

