## Supplementary material for "Effect of Sinotubular Junction Size on TAVR Leaflet Thrombosis: A Fluid-structure Interaction Analysis": Supplementary Document.pdf

### A Particle transport methods

Particle trajectories were predicted by solving Newton's second law and applying a series of forces,

$$\boldsymbol{a}^p = \frac{1}{m^p}\left(\boldsymbol{F}_d + \boldsymbol{F}_g\right)$$

where $\boldsymbol{a}^p$ is the particle acceleration, $m^p = \rho^p V^p$ its mass, $V^p = \frac{1}{6}\pi(d^p)^3$ its volume, and $d^p$ its diameter fixed at $d^p = 3\mu$m, representative of blood platelets. In the following, $\boldsymbol{u}^p$ refers to the particle velocity. The equation for the drag force assumes that the particle has reached its terminal velocity and is given by:

$$\boldsymbol{F}_d = -\frac{\pi}{8}\mu^f d^p C_d \mathrm{Re}\left(\boldsymbol{u}^p - \boldsymbol{u}^f\right)$$

where Re is the particle Reynolds number involving its relative velocity with the fluid:

$$\mathrm{Re} = \frac{\left|\boldsymbol{u}^p - \boldsymbol{u}^f\right| d^p}{\nu^f}$$

with $\nu^f = \mu^f/\rho^f$ the kinematic viscosity. The drag coefficient $C_d$ uses Ganser's formula [1]:

$$\begin{aligned}
C_d &= \frac{24}{\mathrm{Re}\ k_1}\left(1 + 0.1118(\mathrm{Re}\ k_1 k_2)^{0.6567}\right) + \frac{0.4305 k_2}{1 + 3305/(\mathrm{Re}\ k_1 k_2)}, \\
k_1 &= \frac{3}{1 + 2\psi^{-0.5}},\ k_2 = 10^{1.84148(-\log_{10}(\psi))^{0.5743}}, \text{ and} \\
\psi &= \text{ sphericity } (= 1 \text{ for a sphere, as considered here })
\end{aligned}$$

The particles were assumed to be spherical with no interaction with each other. Particle rotation, thermophoretic and Brownian forces were neglected. The particles were sufficiently small, and the suspension sufficiently diluted to neglect their effect on the blood flow (hence the one-way coupling). Buoyancy forces were not considered since the mass density of particles $\rho^p$ were assumed to be the same as blood, that is $\rho^p = \rho^f = 1.1$ g cm$^{-3}$.

### B Construction of stress accumulation PDFs

To obtain the thrombogenic footprint of a specific patient-device setting (as in Figure 7), we select a subset of particles from the total of particles injected into the domain. The subset, which we call $S_{RoI}$, is composed of all the particles whose trajectories at some point passed through the particular Region of Interest (RoI). The fundamental behind this selection criterion is that we intend to study the effect of the passage of platelets through a given RoI on their risk of activation, even though the activation itself may occur elsewhere. In this work, the regions of interest considered are the Sinus of Valsalva and Neosinus. The indices of the considered particles are noted as $p_1, p_2, \dots, p_{N_{RoI}}$, where $N_{RoI}$ is the total number of particles that traversed the region of interest. Therefore, the subsets $S_{SoV}$ and $S_{NS}$ are defined as:

$$\mathcal{S}_{\mathrm{RoI}} = \left\{p_1, p_2, \dots, p_{N_{\mathrm{RoI}}}\right\},\ \forall \mathrm{RoI} \in \{\mathrm{SoV}, \mathrm{NS}\}$$

For each particle $i$, the stress accumulation is recorded at the last registered timestep of its existence (noted as $t_{p_i}^f$ ) and stored as $SA_{p_i} = SA_{p_i}(t_{p_i}^f)$ ). Therefore, the array of stress accumulations for all particles in the $RoI$ subset can be written as:

$$\mathcal{SA}_{\mathrm{RoI}} = \left\{SA_{p_1}, SA_{p_1}, \dots, SA_{p_{N_{\mathrm{RoI}}}}\right\}$$

From the data sample $\mathcal{SA}_{\mathrm{RoI}}$ , a histogram is computed using a logarithmic scale for the bins. Since the Probability Density Function (PDF) cannot be computed exactly from a finite statistical sample, it is estimated using a Kernel Density Estimation (KDE).

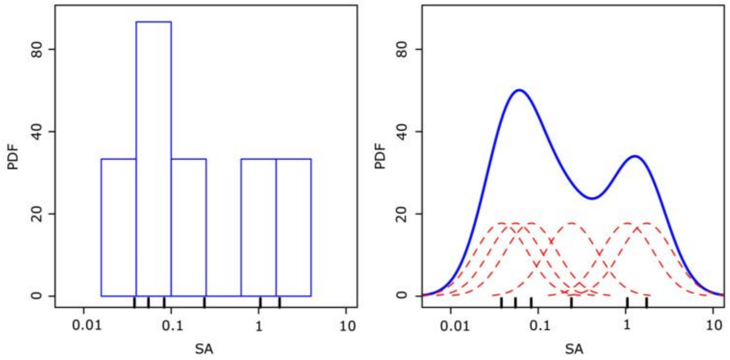


Supplementary Figure 1: Illustration of a Gaussian Kernel Density Estimation (KDE) used to estimate the PDF of a histogram, adapted from [2]. The histogram of the process is shown on the left, while on the right, the dashed red curves represent the individual Gaussian kernels, while the continuous blue curve is the KDE, a linear combination of the kernels.

The KDE is a method to smooth data to infer the underlying PDF from a finite data sample, which is assumed to follow the distribution of some random variable. This process is illustrated in Supplementary Figure 1. Given $(x_1, x_2, \dots, x_n)$ independent and identically distributed samples drawn from a distribution $G, X \sim G$, with unknown PDF $f(x)$, which we want to estimate, the KDE of $f$ is defined as:

$$f_h(x) = \frac{1}{n}\sum_{i=1}^{n} K_h(x - x_i) = \frac{1}{nh}\sum_{i=1}^{n} K\left(\frac{x - x_i}{h}\right).$$

Here $K$ is the kernel $(K > 0), h$ is the bandwidth $(h > 0)$, a smoothing parameter which in this case is chosen as $h = 1$, and $K_h(x) = \frac{1}{h}K\left(\frac{x}{h}\right)$ is the scaled kernel. The kernel may be of one of several types, such as uniform, triangular, Epanechnikov, or Gaussian among others. In this work, we choose the Gaussian kernel for simplicity, which writes:

$$K(x) = \frac{1}{\sqrt{2\pi}\sigma_G} e^{\left(\frac{x-\mu_G}{\sigma_G}\right)^2},$$

where $\mu_G$ is the mean of the distribution $G$, and $\sigma_G^2$ its variance. The extracted histogram is shown to the left of Supplementary Figure 1, with the corresponding PDF estimation to the right. Analogously, this process is carried out for the ensemble of particle Stress Accumulations (SAs) in Figure 7 to compute the SA PDFs in the Sinus of Valsalva and Neosinus.

### C Convergence analysis of stress accumulation PDFs

To assure that the computed SA PDFs are independent of the number of particles injected, a convergence analysis was carried out. More precisely, random ensembles of particles of increasing size were sampled from the full set, in post-process. Then the SA PDFs were computed for these samples of increasing size, as observed in Supplementary Figure 2. The full ensemble contains $1.26 \times 10^6$, $9.96 \times 10^5$, and $1.07 \times 10^6$ particles for the neosinus in the $26$ mm, $30$ mm, and $34$ mm diameter setups respectively. For the SoV, the ensemble contains $1.82 \times 10^6, 1.57 \times 10^5$, and $1.66 \times 10^6$ particles in the $26$ mm, $30$ mm, and $34$ mm diameter setups respectively. Note that the number of particles in all cases is below the total number of injected particles $(2 \times 10^6)$. This is because only the particles that have passed through either one of the RoIs are considered. The three largest samples were observed to differ insignificantly, concluding that the PDFs are independent of the number of injected particles for the full ensemble.

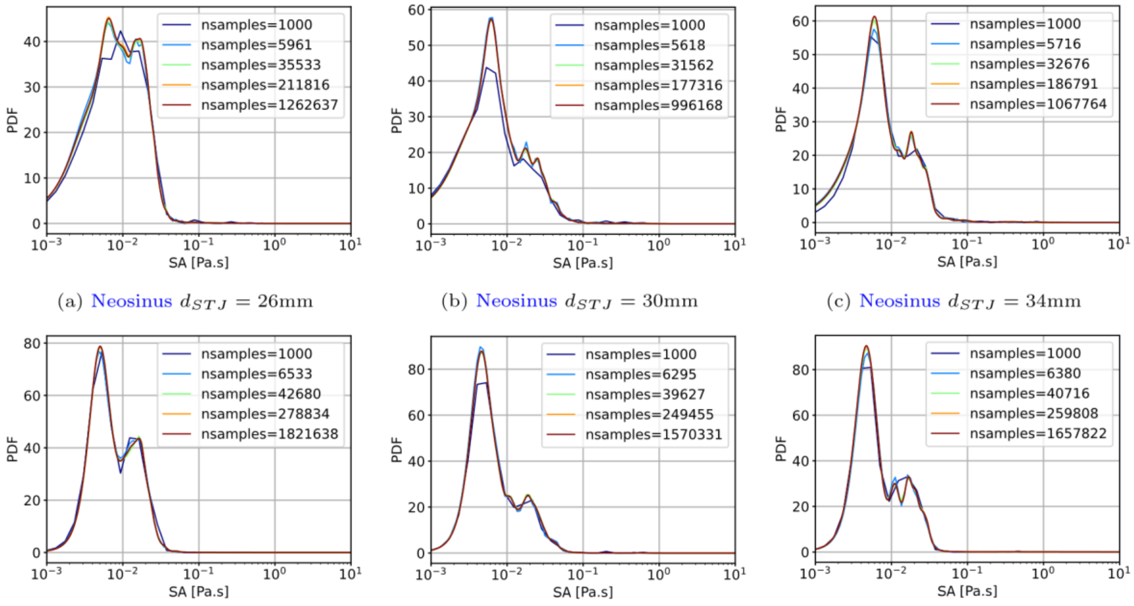


(a) Neosinus $d_{STJ} = 26$mm (b) Neosinus $d_{STJ} = 30$mm (c) Neosinus $d_{STJ} = 34$mm

(d) Sinus of Valsalva $d_{STJ} = 26$mm (e) SoV $d_{STJ} = 30$mm (f) SoV $d_{STJ} = 34$mm

Supplementary Figure 2: Stress accumulation PDFs in the neosinus (top) and sinus of Valsalva (bottom) as a function of particle sample size, for sinotubular junction diameters of: 26mm (left), 30mm (center), and 34mm (right).

### D Comparison between laminar and turbulent model

It is an ongoing subject of discussion if turbulent models are required to model hemodynamics across bioprosthetic and native heart valves, where Reynolds numbers are moderate. Here, we present a comparative analysis between using or not an LES model for the CFD problem. Differences can be observed for the stress accumulation PDFs shown in Supplementary Figure 3, particularly at higher SAs. Here, high SA peaks are augmented for the case with the LES model in both the SoV and neosinus regions of interest. This difference may be caused by the LES model reproducing the average effect of small scales on shear, not accounted for in the laminar model, and thus affecting the resultant stress accumulation in the advected particles. Therefore, in this study, we consider LES modeling for blood flow.

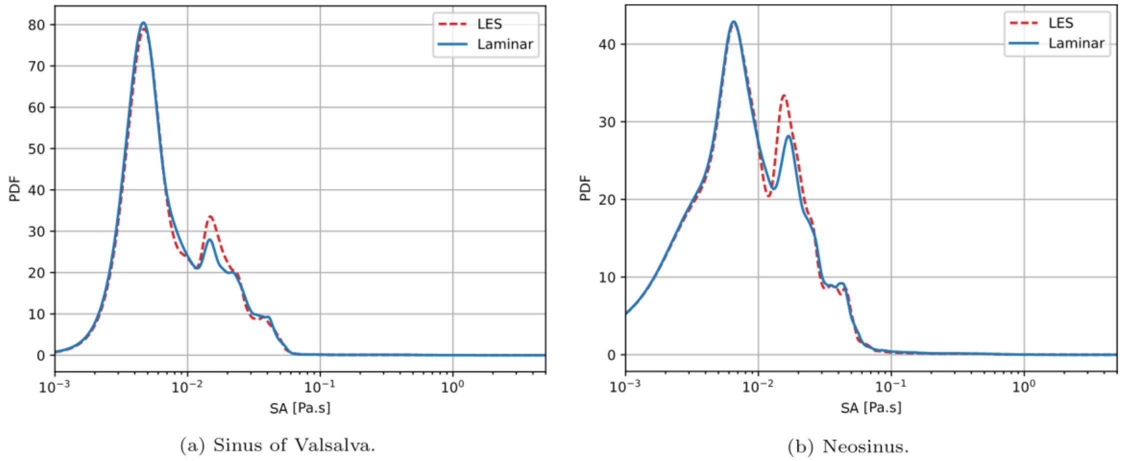


Supplementary Figure 3: Comparison of stress accumulation PDFs in the sinus of Valsalva (left) and neosinus (right), between laminar (blue continuous curve) and LES case (red dashed curve).

### References:


1. Ganser, G., *A rational approach to drag prediction of spherical and nonspherical particles.* Powder Technology, 1993. **77**: p. 143-152.
2. Commons, W., *File:Comparison of 1D histogram and KDE.png* --- *Wikimedia Commons, the free media repository.* 2020.
